# Impact of Temporal Patterns in Working Contacts on Epidemic Spread

**DOI:** 10.64898/2025.12.10.25341893

**Authors:** Aleksandr Bryzgalov, Johannes Ponge, Janik Suer, Tyll Krueger, Beryl Musundi, Chao Xu, Wolfgang Bock, Johannes Horn, Mahreen Kahkashan, André Karch, Julian Patzner, Mirjam Kretzschmar, Rafael Mikolajczyk, OptimAgent Consortium

## Abstract

Most models for infectious disease spread simplify contact heterogeneity by assuming constant rates within a week. However, empirical studies show clear variation, such as reduced workplace contacts on weekends. In this work, we investigate the effects of daily variation in workplace contacts on the spread of respiratory infections using the individual-based framework GEMS (German Epidemic Micro-Simulation System) with a synthetic population of 5 million individuals. We compare a baseline scenario with uniform daily contacts to an alternative with more contacts on workdays and fewer on weekends, keeping weekly totals constant. Simulations reveal that uniform contact rates result in higher prevalence for short latent periods (1–2 days) and reduced one for longer latencies (5–6 days). The effect diminishes for long infectious periods (≥ 7 days) but is more pronounced at lower basic reproduction numbers. Depending on the disease specification and the differences in weekday contacts, the impact of weekday heterogeneity can be very strong. These findings open new possibilities for weekday-dependent mitigation measures. We conclude that weekly contact dynamics should be explicitly incorporated into epidemic models to avoid systematic errors in reproduction number estimation.

## Introduction

Studies have shown that the dynamics of infection spread are specifically influenced by weekends [1, 2, 3]. This includes a clear reduction in workplace contacts and an increase in contacts due to leisure activities, such as social gatherings [4] and increased mobility unrelated to work [5]. The POLYMOD study [6] reported that the number of contacts on weekends was reduced by up to a factor of 1.43 compared with weekdays (see Fig. 1). Similar results were obtained in the COVIMOD study [7].

**Figure 1.**
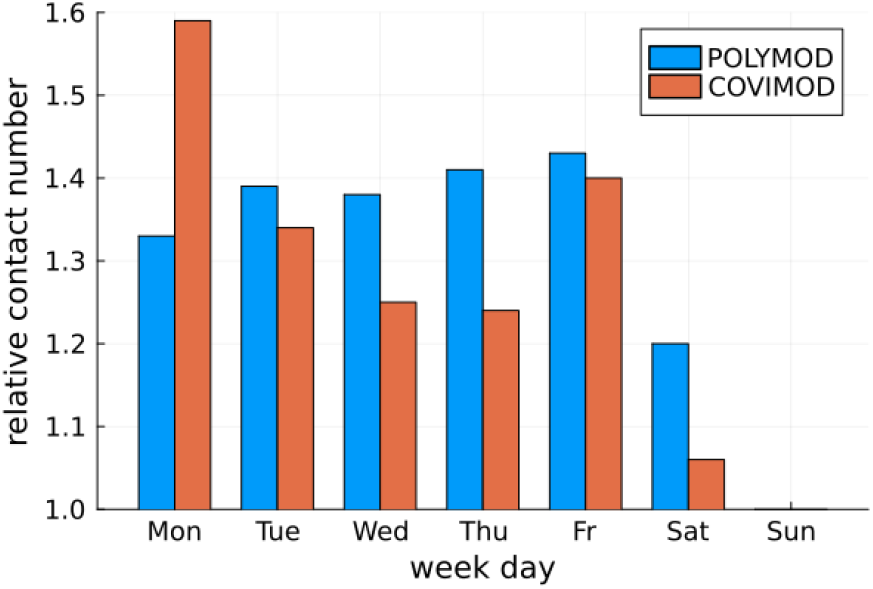
Relative number of contacts depending on weekday from POLYMOD study [6] and COVIMOD Münster study [7]. In both datasets, the minimum number of relative contacts occurs on Sunday and equals 1.

Similar problems appear in areas such as information propagation in social networks [8], resource distribution in supply chains [9], social influence and opinion dynamics [10], telecommunications and network congestion [11], and collaborative workflows in organizations [12]. These problems are typically approached from the perspective of enhancing the spread, seeking optimized solutions to enhance the outcome.

We focus on the consequences of the natural, uneven distribution of contacts throughout the week. How such heterogeneity interacts with the latent period and the duration of infectiousness is crucial for understanding outbreak dynamics. Consider the case where contact rates are very high on Friday but low during the weekend. If both the latent period and the infectious period are short, individuals infected on Friday will have limited opportunities to transmit further, as their infectiousness coincides with days of reduced contact. This example clearly illustrates that epidemic dynamics depend strongly on the weekly structure of contacts and the specific characteristics of the infectious period. In some cases, this can moderate or even halt the spread of infection due to a lack of transmission opportunities. Moreover, spikes in contact rates on particular days may either accelerate or decelerate the spread, depending on how heterogeneity interacts with the latent and infectious periods.

Classical compartment-based modeling using a weekly average contact rate produces similar predictions for the final epidemic size and the timing of the incidence peak, when compared to models that incorporate day-of-week contact patterns [1]. However, the weekday-based model reveals that daily variations in social contacts can generate noticeable fluctuations, or spikes, in the incidence curve of respiratory diseases.

A comparison between predictions using uniform contact rates and those using non-uniform rates could provide valuable insights into the following question: can inaccuracies in the details of contact structure (such as averaging) lead to errors in modeling results? If so, does non-uniformity—or, conversely, uniformity—offer an advantage in controlling the spread of infection? Even when models provide a seemingly good fit to observed data, they may still yield misleading estimates of the reproduction number. In particular, a change in pathogen characteristics—such as a shift in the latent period between variants—can lead to substantial errors if the models rely on assumptions calibrated to earlier strains. As a result, scenario modeling for emerging variants may produce incorrect projections, potentially causing wrong anticipation of outbreak dynamics and the effectiveness of interventions.

In [13], the impact of contact-limiting strategies at workplaces, schools, and high schools was analyzed. More specifically, two approaches were considered: (1) rotating strategies, in which workers are evenly split into two shifts alternating on a daily or weekly basis; and (2) on–off strategies, where the entire group alternates between periods of normal in-person interaction and complete telecommuting. An open question is whether there are alternative ways to balance working time—and thus contact patterns—without reducing the total number of contacts, while still achieving a lower prevalence.

Indeed, certain non-uniform contact restrictions may offer advantages over uniform ones. Depending on epidemiological parameters, targeted restrictions applied on specific days of the week could prove more effective than uniformly distributed restrictions, while requiring fewer overall limitations. Concentrating restrictions on selected days may counterbalance synchronization effects between contact patterns and disease progression, thereby reducing transmission more efficiently. To our knowledge, this aspect has not yet been systematically addressed in the literature and warrants further investigation.

## Methods

As a modeling instrument, we have chosen an individual-based framework called GEMS (German Epidemic Micro-Simulation System) [14]. This framework incorporates the latest insights from the Poland MOCOS model [15], Poland ICM Epidemiological Model[16], and the Austrian full-scale agent-based model [17].

### GEMS description

GEMS individual-based infectious disease modeling framework. It simulates the spread of respiratory pathogens within a synthetic population in discrete steps (e.g., days). GEMS operates using two core entities: individuals and settings. Each individual is assigned a set of attributes. Some attributes remain fixed throughout the simulation (e.g., ID number, age, gender, household ID, workplace ID), while others may change over time (e.g., susceptibility, disease state, symptom category).

Settings represent the environments where individuals interact. In this study, we used the following settings: household, workplace, school class (including kindergartens for children under six), and a global setting. The global setting allows any individual to come into contact with any other, unlike the other settings, where contacts are restricted to the “residents” of that specific environment.

Based on age, individuals are either assigned to a school/kindergarten or to a workplace. Pensioners neither study nor work and therefore do not have contacts in these specific settings.

Together, all individuals constitute the simulation population.

In the configuration used in this work, each infected individual progresses through the standard SEIR disease states: Susceptible (S), Exposed (E), Infectious (I), and Recovered (R). To simplify the model, reinfection (R → S transition) is not included.

At the start of each simulation run, a randomly selected subset of individuals is initialized in the exposed (E) state. A random day in the exposure interval is used. Susceptible individuals can become infected through contact with infectious individuals, with the transmission probability depending on the setting in which the contact occurs. New individuals are the earliest to be infected on the next day, due to the step-by-step nature of the process.

We focused exclusively on the setting (e.g., place) of a contact as the determining factor for transmission.

### Population

The population was synthetically generated based on predefined distributions of households, workplaces, and schools (including kindergartens). This flexible structure allowed us to isolate and assess the effects of weekday variations. In our study, we used a synthetic population of 5,000,000 individuals. The following mean sizes were specified for the settings:

1. Households: mean size of 2, consistent with the average household size in Germany [18] (see also Fig. 2 (a));
2. Workplaces: sizes ranging from 2 to 100, depending on the specific task or simulation scenario;
3. School classes and kindergarten groups: fixed at a mean size of 20.

**Figure 2.**
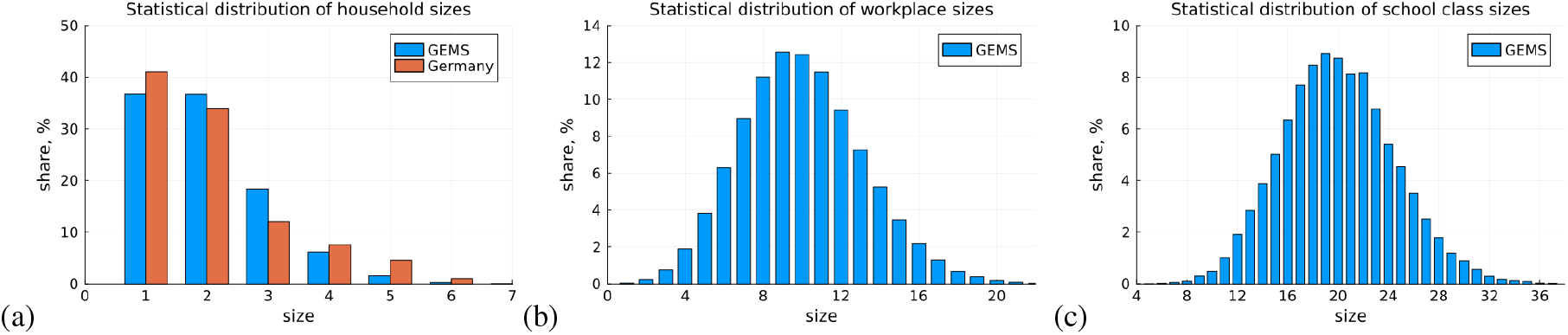
Distribution of households (a), workplaces (b) and school classes (c), generated by GEMS synthetically. In Fig. (a) German distribution of households [18] is shown additionally.

In all cases, the size distributions were zero-truncated Poisson-distributed. An example of these distributions is shown in Fig. 2.

The age distribution was: 18.5% with 0 - 17 age (contacts in school and kindergarten), 66.5% with 18-64 age (working population) and 15% with 65 + age (elderly, not working population). The initial infected share was 0.1% of the population (5000 individuals).

### Disease progression parameters and infectivity per contact

Infected individuals progress through the SEIR disease states over time. To simplify the analysis, we define the latent period as the time between exposure and the onset of symptomatic infectiousness, effectively representing the difference between the incubation period and the duration of asymptomatic infectiousness (see also Tab. 1).

**Table 1.** Description of disease progression from the [19] and input parameters for the pathogen that were used in simulations. Since there is no direct correspondence, we considered the difference between the onset of symptoms and the infectious offset as the latent period. For an infectious offset of 0, the latent period *L* corresponds to the onset of symptoms, and the infectious period *I* corresponds to the time of recovery. In most simulations, we used exact values for *L* and *I* to have a pure impact.

| Different states of infection | Description from [19] | GEMS parameter name | Value |
| --- | --- | --- | --- |
| latent period | On average, viral shedding was detected 1 day after inoculation | *infectious offset* determines if an individual becomes infectious before becoming symptomatic | 0 |
| incubation period | Systemic symptoms (fever, muscle aches, fatigue, headache) peaked earlier, by day 2 after inoculation | *onset of symptoms* is the time when symptoms start | 1 |
| infectious period | The mean duration of viral shedding was 4.80 days | *time to recovery* is the time to recover from an infection as an increment from the onset of symptoms | 5 |

For the pathogen model, we used influenza A/H1N1, with parameter values obtained from [19]. Additionally, we include the Fig. 3 for a clear explanation of these parameters.

**Figure 3.**
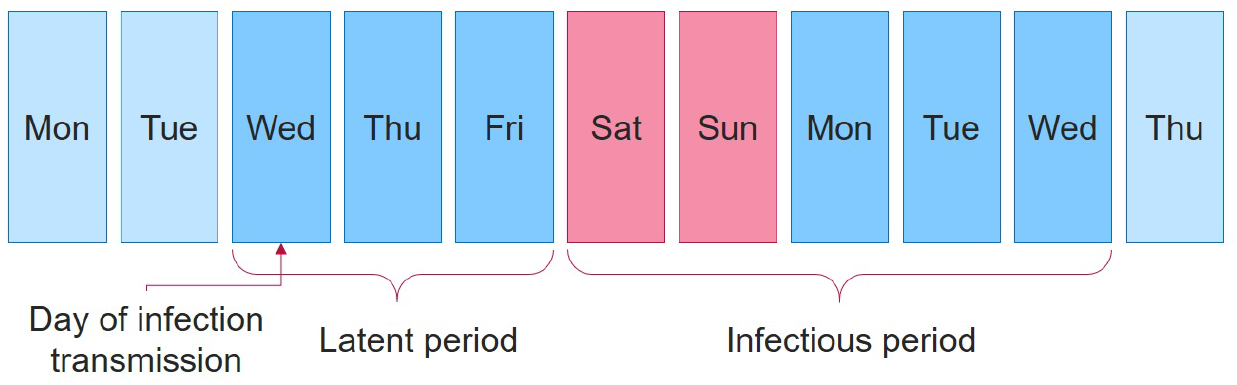
Example of the pathogen parameter relation. In our simulations, the day of infection transmission is considered to be fully included in the latent period

Moreover, the chosen values for the infectious and latent periods are appropriate for SARS-CoV-2 infection. Although the overall infectious period exceeds 7 days, the effective duration during which individuals are outside the household—and thus able to transmit the virus in the community—is considerably shorter, as illness typically confines them to the home.

Another aspect we considered in our analysis is the variation in infectivity across different settings. Broadly, we distinguish three main settings of transmission: households, workplaces and schools/kindergartens, and other locations (global setting). Contacts within households and workplaces (including schools and kindergartens) tend to be more stable, as individuals typically interact with the same group daily. In contrast, contacts in other settings are more variable. We assumed higher infectivity for contacts within households and workplaces compared to those in the global setting [20].

Model calibration was performed for the baseline scenario to estimate transmission rates per contact. The target distribution of infections was as follows: one-third within households, one-third within workplaces and schools (including kindergartens), and one-third in other locations (global setting). This distribution represents a compromise between observed infection data [20, 21, 22] and the relative number of contacts occurring in these settings [6].

As outcomes, we considered the prevalence during the simulation period.

### Scenarios

As a baseline scenario, we considered a fixed number of contacts each day. In an alternative scenario, we redistributed the contacts, reducing them on weekends for children and the working population (only for workplaces, schools and kindergartens). The reasoning behind this is clear: usually, people do not work during weekends, and children do not go to school or kindergarten. The total number of contacts was the same in both scenarios.

In the baseline scenario, contact rates are independent of the weekday. The total numbers for a week are as follows: 3 × 7 = 21 contacts at the workplace, 6 × 7 = 42 contacts at school (including kindergarten), and 5 × 7 = 35 contacts in the global setting. See also Tab. 2.

**Table 2.** The number of daily contacts and the transmission probability per contact. The global setting refers to all locations except those specified in the table columns. This combination of values gives an effective reproduction number of approximately 1.2.

| Setting | Household | Workplace | School class/<br>Kinder-garten | Global setting |
| --- | --- | --- | --- | --- |
| Number of contacts per day<br>$n$ | Size - 1 | 3 | 6 | 5 |
| Transmission probability per<br>contact $p$ | 0.0600 | 0.0300 | 0.0300 | 0.0132 |

In the alternative scenario, we redistributed the contacts across weekdays while keeping the total number of contacts for each setting unchanged. See Tab. 3.

**Table 3.** The mean contact rates in alternative scenario.

| Setting | Workplace |  | School class/<br>Kindergarten |  | Global Setting |  |
| --- | --- | --- | --- | --- | --- | --- |
| Week day | Workday | Weekend | Workday | Weekend | Workday | Weekend |
| Mean number of contacts<br>per day $n$ | 4.2 | 0 | 8.4 | 0 | 5 | 5 |

We used sampling with replacement. This means that the probability of being infected in a smaller workplace is higher. Most simulations were done with a workplace size of 10.

### Estimation of expected number of infections by spectral radius

Previously, we have spoken only about the simulation opportunities, which themselves should be verified and validated. Such work was partially done, see, for example, [14]. To close this gap, we introduce an analytical method. It can be used as a preliminary evaluation to quickly assess the effects of temporal contact structures. Specifically, we can compare a uniform contact distribution with a non-uniform one—for example, cases with increased contact rates on working days—without running full simulations. However, these evaluations rely on several assumptions: the method provides the estimation for a single population group, it does not take into account correctly the contacts in the household, and the workplace size should be sufficiently large. This also implies that GEMS simulations provide more precise results.

However, since we consider the changes in contacts in the workplace, the estimations done by the analytical method for the working adults form a qualitative impression of what simulation results we expect for given daily contact rates.

To calculate the expected number of secondary infections *N*_*total*_ during the infectious period *I*, we use the spectral radius *ρ*. It is well known that the expected number of secondary cases produced by a typical infected individual during their entire infectious period in a completely susceptible population is mathematically defined as the dominant eigenvalue (i.e., the spectral radius) of a positive linear operator [23].

We classify individuals into types according to the weekday on which they become infected. The matrix

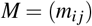

specifies the expected number of type *j* individuals generated by a type *i* individual during its infectious period. This does not depend on type, since types are equally created and only impact the number of secondary cases generated by an individual.

Thus, we can construct the matrix *M* based on the contact structures shown in subsection Scenarios. Generally, each row of *M* represents the number of infectious contacts, depending on the day of transmission, the latent period *L*, and the infectious period *I*. For example, if transmission occurred on a Monday with a latent period *L* = 2 days and an infectious period *I* = 5 days, the corresponding row would look as follows:

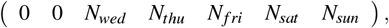

where *N*_*wed*_,*N*_*thu*_, *N*_*f ri*_, *N*_*sat*_, *N*_*sun*_ are the corresponding number of infectious contacts on a certain weekday.

To calculate the infectious contacts on a given day *N* we multiply the infectivity in a setting *p* by the corresponding number of contacts *n*:

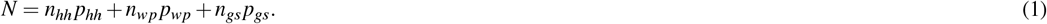

The index *hh* is household, the index *wp* is the workplace and the index *gs* means global setting. Obviously, the values of *N* will not differ for the uniform distribution of contact rates and infectious period *≤I* 7 days. For a longer infectious periods, a certain weekday can be covered twice or more times, which results in a proportional increase of *N*. Also, we notice that *I* = 7; 14; 21; …will not give any differences in *ρ* between scenarios, since the number of weekend days covered will be the same.

In the simplest case, we consider that the population is highly mixed and the contacts take place only in the global setting:

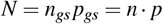

For a simplified explanation, we assign numbers, which give a similar value of *R*_0_ to the ones that include household contacts and working contacts. In baseline scenario every individual has *n*_uniform_ = 3 contacts per day. In an alternative scenario, on working days the contact rate is *n*_work_ = 4.2 and on weekends *n*_we_ = 0. Infectivity per contact is *p* = 0.075 to get the reproduction number greater than one. The corresponding numbers of infectious contacts will be: *N*_uni_ = *p n*_uniform_; *N*_work_ = *p n*_work_ and *N*_we_ = *p n*_we_.

Let’s consider the infectious period *I* = 6 days and the latent period *L* = 1 day. Thus, we can construct the matrices *M*_*base*_(baseline) and *M*_*alt*_ (alternative) of next-generation infections (see [23]) that can be written as:

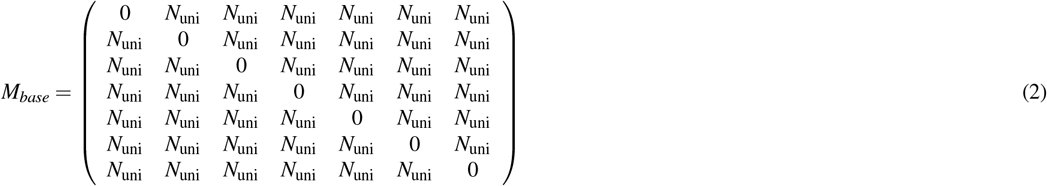

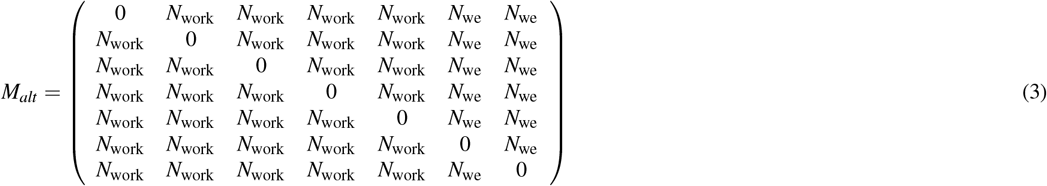

Columns reflect the weekday and the rows display the day of infection transmission (zeros in matrices).

Then, the spectral radius is:

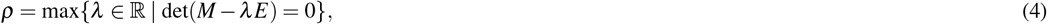

where *E* is identity matrix and *λ* is eigenvalue.

Moreover, the prevalence *a*_*total*_ can be obtained. According to the general theory of branching processes [24], the left eigenvector

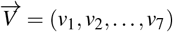

of *m*_*i j*_, normalized such that ∑ *v*_*i*_ = 1, gives the relative share of type *i* occurring in the asymptotics of the branching process. Furthermore,

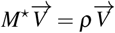

with *ρ* being a spectral radius. We claim that the relative proportions of the different types of individuals are still given by 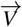, since the types are not fixed a priori and no additional heterogeneity is assumed in the population. Even in the presence of a saturation effect, each individual has access to the same “pool of resources”, which does not alter the spectral radius or the corresponding eigenvector.

Let us consider a test individual drawn from a sufficiently large population with a given infected fraction. Each individual generates a Poisson-distributed in-degree. Thus, the likelihood that the test individual becomes infected, denoted by *a*_*total*_, is given by:

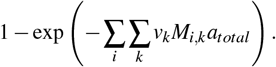

Then

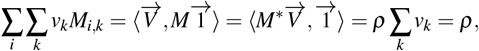

where 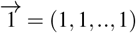. Thus, we obtain for the prevalence *a*_*total*_ the following formula

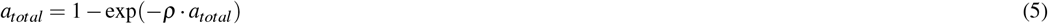

Using the values mentioned above *n*_uniform_, *n*_work_, *n*_we_ and *p*, we calculate the spectral radii as *ρ*_*baseline*_ = 1.26 and *ρ*_*alternative*_ = 1.35 and corresponding *a*_*total*_.

Additionally, we provide an estimation from the classical SEIR compartmental model [25] using the *R*_0_ = *N*_uni_*I* and the results based on GEMS simulations. The comparison is shown in the Tab. 4.

**Table 4.** Comparison of estimations of prevalence (%) for a highly mixed population. The results are obtained by analytical formula (5), SEIR equation-based model and individual-based framework GEMS.

|  | formula (5) | SEIR equation-based model<br>[25] | GEMS |
| --- | --- | --- | --- |
| baseline | 46.9 | 47.1 | 46.9 |
| alternative | 38.2 | - | 38.4 |

The generalization of matrices (2) and (3) for the different population groups (e.g., children, working adults, and the elderly non-working population) is not included in this work, nor is the extended form of formula (5). Further investigation is needed for a more precise assessment of household transmission in the calculation of the spectral radius.

However, even assessing a single dominant population group can provide an upper bound for *R*_0_. We performed this estimation for working adults, assuming that each working individual has one household contact. This results in the number of infectious contacts on a working day being: *N*_work_ = *n*_*hh*_ *p*_*hh*_ + *n*_*wp*_ *p*_*wp*_ + *n*_*gs*_ *p*_*gs*_ = 1 0.06 + 4.2 0.03 + 5 0.0132 = 0.252 and on weekend day *N*_we_ = 0.126.

Spectral radius estimations are included in the Results section and in the Appendices.

## Results

In this section, we analyze the effect on the prevalence of the redistribution of contacts combined with variations of the latent period, infectious period, workplace size and infectivity per contact. Also, we provide estimations based on the spectral radius (see the previous section).

For each workplace size within a given scenario, we conducted 100 simulations, calculating the mean values and confidence intervals for the results. The simulation time was usually set to 500 days.

### Variation of latent period

In this section, we do not restrict ourselves to a specific type of pathogen and instead study the influence of the latent period on the results.

The latent period typically lasts several days before the infectious period begins. In the alternative scenario, we observe more infections than in the baseline scenario for a 5-day latent period (Fig. 4 right). For a 1-day latent period, the pattern is reversed (Fig. 4 left). This outcome was expected based on spectral radius calculations. For more details, see the table in Appendix 1.

**Figure 4.**
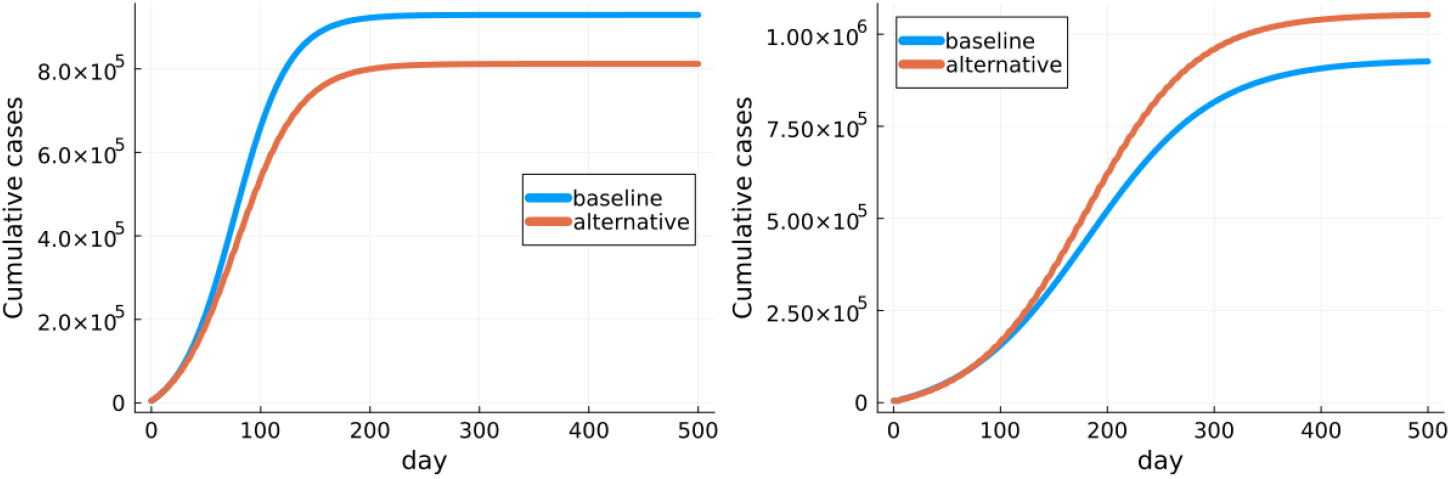
Prevalence for different scenarios. Left: latent period is 1 day and simulation time is 500 days; prevalences are: baseline (18.58*±*0.15)%, alternative (16.24*±*0.16)%. Right: latent period is 5 days and simulation time is 500 days; prevalences are: baseline (18.52*±*0.15)%, alternative (21.04*±*0.16)%

In the simulations shown in Fig. 4, we used exact values for the latent period.

### Variation of workplace size

The dependence on workplace size is shown in Tab. 5. In the baseline scenario, we fixed the number of contacts at 3, and in the alternative scenario, at 4.2 during working days. For schools, these rates are 6 and 8.4 correspondingly. As a result, there is a natural moderating effect for infection spread in small workplaces (i.e., those with fewer than 5 people). For very large workplaces (i.e., those with more than 40 people), the increase in infections reaches a saturation point. This outcome is expected, as the number of possible contacts in such large workplaces is high enough to allow the infection to spread more widely.

**Table 5.**
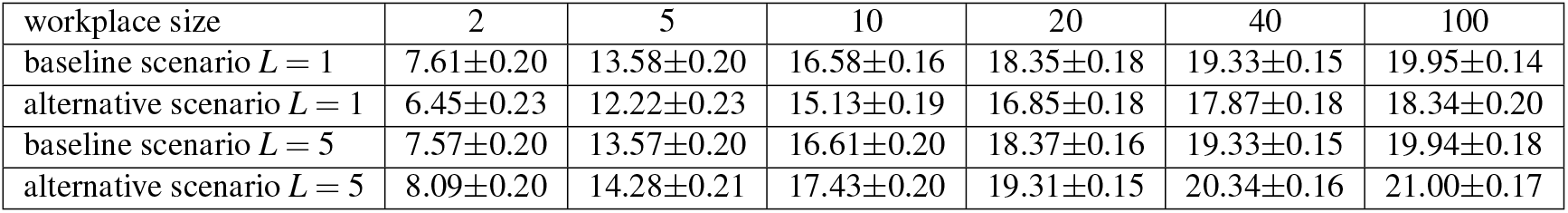
Prevalence (%) for the baseline and alternative scenarios for different workplace sizes. The sizes of other settings, including schools, remain unchanged. The transmission probability per contact and contact rates are taken from Tab. 2, the values of the latent period *L* are exact and equal to 1 and 5, the mean value of the infectious period *I* is 5, and the distribution is zero-truncated Poisson.

| workplace size | 2 | 5 | 10 | 20 | 40 | 100 |
| --- | --- | --- | --- | --- | --- | --- |
| baseline scenario $L = 1$ | $7.61 \pm 0.20$ | $13.58 \pm 0.20$ | $16.58 \pm 0.16$ | $18.35 \pm 0.18$ | $19.33 \pm 0.15$ | $19.95 \pm 0.14$ |
| alternative scenario $L = 1$ | $6.45 \pm 0.23$ | $12.22 \pm 0.23$ | $15.13 \pm 0.19$ | $16.85 \pm 0.18$ | $17.87 \pm 0.18$ | $18.34 \pm 0.20$ |
| baseline scenario $L = 5$ | $7.57 \pm 0.20$ | $13.57 \pm 0.20$ | $16.61 \pm 0.20$ | $18.37 \pm 0.16$ | $19.33 \pm 0.15$ | $19.94 \pm 0.18$ |
| alternative scenario $L = 5$ | $8.09 \pm 0.20$ | $14.28 \pm 0.21$ | $17.43 \pm 0.20$ | $19.31 \pm 0.15$ | $20.34 \pm 0.16$ | $21.00 \pm 0.17$ |

The effect of the latent period also occurs: in the baseline scenario, higher prevalences (compared to the alternative scenario) are obtained for each workplace size when *L* = 1, and lower prevalences when *L* = 5.

We want to emphasize that the contacts were redistributed throughout the week only at workplaces and schools (kindergartens). Contacts in households and in the global setting remained fixed.

### Variation of infectivity

During the simulations, we observed a stronger effect of the redistribution of contacts in scenarios with a smaller prevalence. See Table 6.

**Table 6.**
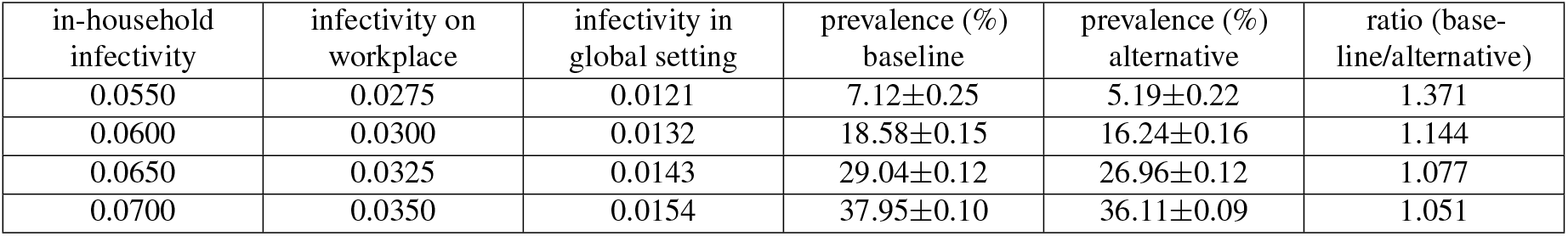
Comparison of the workday/ weekend effect for the different infectivity. The results are given for the case of workplace size 10. Contacts in the alternative scenario were changed according to the Tab. 3. Infectivity in settings (home, work and global setting) was changed proportionally to save the corresponding infection distribution in settings.

From the table, it is evident that cases with lower infectivity are more sensitive to changes in the contact rate distribution. A similar effect can be achieved by varying the infectious period while keeping the infectivity per contact constant. Increasing the number of infectious days leads to more infections.

### Interventions, based on contact redistribution

Redistribution of contacts can be used as an intervention strategy. We suggest considering the addition of a weekend day and a shortened week structure with two working days and two days off, while maintaining the same total number of weekly contacts. The effects are highly dependent on the latent and infectious periods; therefore, the choice of strategy should be more closely aligned with the characteristics of the pathogen.

#### Additional weekend day with a proportional increase of working time

We propose the following contact rate redistribution (Tab.7).

The idea is that the working time should be proportionally increased to equalize the economic effect (ignoring logistics restructuring of organisations). Of course, here our interest is to compare the scenario of 2 weekend days with 3 weekend days. And we still have the same number of contacts.

**Table 7.** Contact rates in scenario with 3 weekend day working week. Corresponding weekly contact rates are: 21 contacts in the workplace and 42 contacts in the school.

| Setting | Workplace |  | School class/ Kindergarten |  | Global Setting |  |
| --- | --- | --- | --- | --- | --- | --- |
| Week day | Mon-Thu | Fri-Sun | Mon-Thu | Fri-Sun | Mon-Thu | Fri-Sun |
| Mean number of contacts per day | 5.25 | 0 | 10.5 | 0 | 5 | 5 |

We provide a comparison of infectious contact numbers in Fig. 5 based on spectral radius calculations for the one population group - working adults (see table in Appendix 2). This means these are the estimations that give a qualitative understanding, what the prevalence change could be in case of redistribution of contacts. The figures have been obtained using the tables from Appendices 1-3.

**Figure 5.**
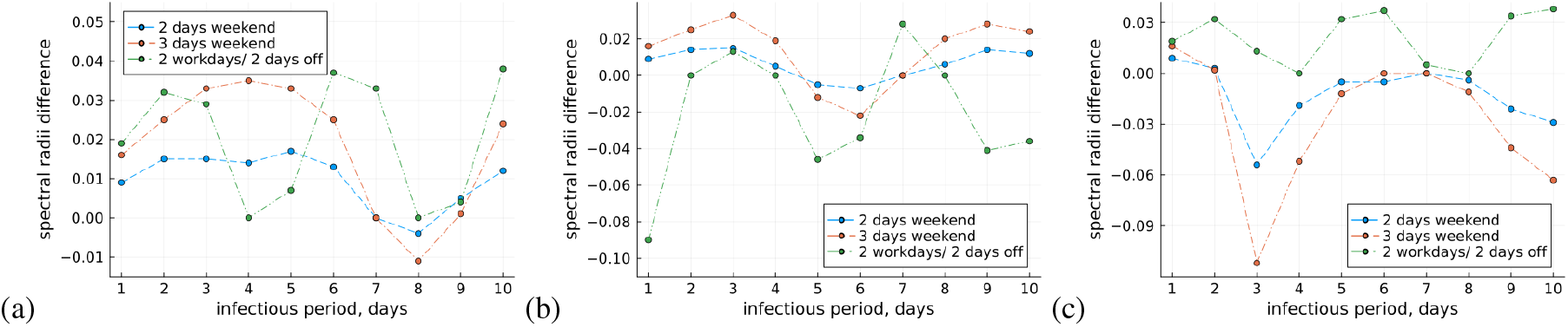
Spectral radii difference *ρ*_*baseline*_*− ρ*_*alternative*_. Alternative scenarios are: 2 days weekend - normal week with 2 weekend days, 3 days weekend - week with 3 weekend days, 2 workdays/2 days off - 2 working days, 2 days off. Weekly contacts are the same in all scenarios. (a) latent period is 1 day; (b) latent period is 4 days and (c) latent period is 6 days. The used infectivity values are: *p*_*hh*_ = 0.06, *p*_*wp*_ = 0.03 and *p*_*gs*_ = 0.0132. This means for the values of *I <* 5 the corresponding *ρ*_*baseline*_ *<* 1

Based on the analysis of spectral radii calculation results, we choose specific combinations of infectious period *I* and latent period *L* for simulations. Infectivity per contact has also been updated to keep the *R*_0_ *>* 1. We summarize the GEMS simulation results in the Tab. 8.

**Table 8.** Comparison of the simulation results for 2 days weekend week, 3 days weekend week and 2 working days/ 2 days off week. Also, we provide data for the baseline scenario (with uniform contact rates). Case 1: *L* = 1 day, *I* = 7 days, *p*_*hh*_ = 0.04, *p*_*wp*_ = 0.02 and *p*_*gs*_ = 0.0088. The results for 2 days and for 3 days weekend are not different from the baseline scenario, since the infectious period is 7 days. Case 2: *L* = 1 day, *I* = 5 days, *p*_*hh*_ = 0.06, *p*_*wp*_ = 0.03 and *p*_*gs*_ = 0.0132. Case 3: *L* = 6 day, *I* = 3 days, *p*_*hh*_ = 0.09, *p*_*wp*_ = 0.045 and *p*_*gs*_ = 0.0198. Exact values without distributions for *L* and *I* have been used.

| week structure | Case 1 | Case 2 | Case 3 |
| --- | --- | --- | --- |
| 2 days weekend | - | 16.24±0.16 | 14.46±0.20 |
| 3 days weekend | - | 13.82±0.20 | 24.30±0.12 |
| 2 w.d./2 days off | 6.10±0.24 | 17.64±0.18 | 2.79±0.31 |
| baseline | 9.16±0.24 | 18.58±0.15 | 5.08±0.18 |

**Table 9.** Contact rates in scenario with 2 working days / 2 days off working week structure. To obtain the numbers, we use the following: 21 contacts should be in a 7-day period in the workplace and 42 contacts in a school; the mean number of weekend days in a 7-day period is 3.5.

| Setting | Workplace |  | School class/<br>Kindergarten |  | Global Setting |  |
| --- | --- | --- | --- | --- | --- | --- |
|  | Every 1-2<br>day | Every 3-4<br>day | Every 1-2<br>day | Every 3-4<br>day | Every 1-2<br>day | Every 3-4<br>day |
| Mean number of<br>contacts per day | 6 | 0 | 12 | 0 | 5 | 5 |

#### 2 working days / 2 days off working week structure

Some types of work require round-the-clock coverage, rotating shifts, or continuous operations, including nights and weekends. In such cases, shifts can be organized as 2 working days followed by 2 days off (see Tab. 9).

The estimations of spectral radii are shown in Appendix 3. The “week” in this case is shorter (4 days in total). So the durations of the infectious period in days that are divisible by 4 with no remainder do not produce any differences between scenarios.

For a chosen simulation results see Tab. 8. For example, the 2 working days / 2 days off working week structure can provide 1.5 times less prevalence (6% versus 9% in baseline) in the case of *L* = 1 and *I* = 7.

## Discussion

We conclude that a non-uniform distribution of contacts throughout the week can lead to either a positive or a negative outcome. This effect arises from the discrepancy between the duration of the infectious period and the structure of the working week. Additionally, we conclude that this effect should be included in any agent-based model simulating daily contacts or a compartmental model as well.

The strongest effect occurs when the effective reproduction number is close to 1 and the infectious period is short (< 7 days). Moreover, the difference between contact rates on working days and weekend days should be significant (for example, 4.2 mean contacts on working days versus 0 mean contacts on weekend days at work and school).

Specific combinations of latent period, infectious period, and infectivity per contact can cause completely different dynamics in baseline and alternative scenarios involving phase transitions (see Tab. 8). We do not provide simulation examples for all cases within this work; however, some estimations can be given by spectral radius calculation. Please see the Appendices 1-3 for that and Fig. 5.

The key factors associated with the redistribution of contacts are summarized in Tab. 10.

**Table 10.** Summary of the effects produced by the given factors combined with contact redistribution.

| Factor (parameter) | Range of values | Comment |
| --- | --- | --- |
| latent period | 1-10 days | defines how many weekend days will be included in the infectious period, which forms the key difference between the scenarios. |
| infectious period | 1-10 days | the 3-4 day infectious period introduces more heterogeneity, resulting in differences between the alternative and baseline scenarios; 7-day infectious period resulted in no differences |
| workplace size | 2-100 sizes | saves the deviation between scenarios; for larger workplaces the relative difference is smaller |
| difference between numbers of contacts in a given setting in workday and weekday | 0; 3; 4.2; 5.25<br>contacts in workplace<br>0; 6; 8.4; 10.5<br>contacts in school | as a higher difference in contacts in working and weekend days as stronger could be the effect of redistribution |
| infectivity per contact | 0.055 - 0.070<br>in-household<br>infectivity | higher infectivity increases outcome; simulations with smaller infectivity are more dependent on contact distribution |

### Impact of working contacts on prevalence

The adult population share is 66.5% and 18.5% of children in our simulations. But, of course, the changes that we applied for working contacts in the alternative scenario do not affect the non-working part of the population (approximately 15%). Moreover, we must account for other contacts (household or global setting), which remain the same and can support the spread after the spike.

We also conclude that the effect of workplace size is much stronger than the redistribution of contacts during the week (Tab. 5).

### Adaptation of working week structure to the type of the pathogen/ respiratory infection disease

Additionally, we have considered scenarios that can support a specific kind of interventions, balancing the working time. The general idea is to leverage the advantages of contact redistribution, depending on the pathogen or disease type.

For the latent period *L* = 1 day and infectious period *I <* 7 days (Fig. 5 (a)), the additional weekend day is advantageous. Latent period *L* = 4 days and *I* of 5 or 6 days (Fig. 5 (b)) give the estimations that it is preferable to keep a normal week with 2 weekend days, since the additional weekend day with working time saving may increase the prevalence. For the latent period *L* = 6 days (Fig. 5 (c)), redistribution of having two or three weekend days increases the prevalence in most cases. Very dangerous case of *I* = 3 days that can boost the infection spread (see Case 3 in Tab. 8). A good type of intervention here would be alternating two working days with two days off.

The provided method for estimating the spectral radius can be used as a preliminary qualitative result.

### Possible generalizations

We identify several directions for future work:

- We employed a completely synthetic population, which allowed us to treat workplace size as a tunable parameter. A promising avenue for future analysis is the comparison of detailed, fitted simulations of the temporal dynamics of the reproduction number between models that explicitly incorporate dynamic contact rates and those that instead rely on averaged contact patterns.
- >We did not consider a U-shaped distribution of workplace sizes. Nevertheless, we expect similar qualitative effects, given the observed correlation between workplace size and infection outcomes.
- >Reinfections (i.e., transitions from recovered to susceptible) were not included, as they were not expected to have a major impact. In most simulations, prevalence after 300 days closely matched the values observed at 500 days. Naturally, reinfection dynamics should be incorporated in future work to investigate the endemic equilibrium state.
- >Variation in household sizes could also be explored.
- >The influence of the distributional shape of the latent and infectious periods merits further study. In this work, we primarily considered fixed values and employed a zero-truncated Poisson distribution in simulations with workplace size variation. Alternative distributions may affect the results in different ways.
- >Regional heterogeneity represents another important factor. For instance, analyzing infection spread in bordering regions with differing working-week structures would provide an interesting case study.

Real data show more complicated patterns of contact distribution throughout the week [6, 7, 26]. The actual proportions of contact redistribution could differ from the scenarios considered here and may yield more significant effects.

Moreover, some specific aspects can be studied additionally, like vaccination coverage effects for different age groups or investigation of occupations’ effect (see, for example [27, 28]).

Also, we do not consider the aspects relating to the asymptomatic disease progression, which was switched off (the *infectious offset*= 0). However, this factor makes it difficult for authorities to take effective control measures [29]. This question becomes more complicated when taking into account the different susceptibility between different age groups [30]. Further study should include the possibility of self-isolation for correct estimations regarding the coupling of these effects with the redistribution of contacts.

## Data Availability

All data produced in the present study are available upon reasonable request to the authors

https://github.com/IMMIDD/GEMS

## Acknowledgements

We acknowledge the contributions of the OptimAgent consortium and its members for their support and collaboration in this research: Alexander Kuhlmann (Institute for Social Medicine and Epidemiology, University of Lübeck, Germany); Myka Sarajan, Carla Hartmann (Institute for Medical Epidemiology, Biometrics and Informatics (IMEBI), Interdisciplinary Center for Health Sciences, Medical School of the Martin-Luther University Halle-Wittenberg, Halle (Saale), Germany); Hannah Derwanz; (Faculty of Medicine, Martin Luther University of Halle-Wittenberg, Halle (Saale), Germany); Bernd Hellingrath, Johannes Ponge (Chair for Information Systems and Supply Chain Management, University of Münster, Münster, Germany); Veronika K Jaeger, Huynh Thi Phuong (Institute of Epidemiology and Social Medicine, University of Münster, Münster, Germany) Vitaly Belik, Andrzej K. Jarynowski, Marlli Zambrano (System Modeling Group, Institute of Veterinary Epidemiology and Biostatistics, Freie Universität Berlin, Germany) Steven Schulz, Richard Pastor, Alejandra Rincon, Ashish Thampi (Machine Learning and Health Unit, Department of Engineering, NET CHECK GmbH, Berlin, Germany) André Calero Valdez, Lilian Kojan (Institute of Multimedia and Interactive Systems, University of Lübeck, Lübeck, Germany); Markus Scholz (Institute for Medical Informatics, Statistics and Epidemiology, University of Leipzig, Leipzig, Germany); Jan Pablo Burgard, Soheil Shams, João Vitor Pamplona (Department of Economic and Social Statistics, Trier University, Trier, Germany); Wolfgang Bock (Department of Mathematics, Linnaeus University, Växjö, Sweden); Lukas Bayer, Sudarshan Tiwari (Department of Mathematics, RPTU Kaiserslautern, Kaiserslautern, Germany); Berit Lange, Isti Rodiah (Department of Epidemiology, Helmholtz Centre for Infection Research, Brunswick, Germany); Wolfgang Greiner, Maren Steinmann, Sebastian Gruhn (Department of Health Economics and Health Care Management, School of Public Health, Bielefeld University, Bielefeld, Germany); Uwe Siebert, Beate Jahn (Institute of Public Health, Medical Decision Making and Health Technology Assessment Department of Public Health, Health Services Research and Health Technology Assessment UMIT TIROL – University for Health Sciences and Technology, Hall. i.T., Austria).

## Author contributions statement

**Aleksandr Bryzgalov**: conceptualization; design and formalization of theoretical aspects; data analysis; model development and implementation; simulations; analysis of the results; text writing. **Johannes Ponge**: model development and implementation. **Janik Suer**: model development and implementation. **Tyll Krueger**: model development, conceptualization, design and formalization of theoretical aspects; analysis of the results; text writing. **Beryl Musundi**: conceptualization, data analysis; simulations. **Chao Xu**: conceptualization, data analysis. **Wolfgang Bock**: model development, conceptualization. **Johannes Horn**: model development, conceptualization, data analysis. **Mahreen Kahkashan**: conceptualization, data analysis. **André Karch**: analysis of the results. **Julian Patzner**: model implementation. **Mirjam Kretzschmar**: conceptualization; design and formalization of theoretical aspects; analysis of the results. **Rafael Mikolajczyk**: conceptualization; design and formalization of theoretical aspects.

All authors reviewed the manuscript.

## Additional information

### Declaration of generative AI use

The authors used generative AI to enhance the clarity and grammar of this manuscript. However, the manuscript was carefully reviewed by all authors to ensure accuracy and originality.

### Ethical approval

We certify that this manuscript is original and has not been published and will not be submitted elsewhere for publication while being considered by Scientific Reports. No data have been fabricated or manipulated (including images) to support our conclusions. No data, text, or theories by others are presented as if they were our own. And authors whose names appear on the submission have contributed sufficiently to the scientific work and therefore share collective responsibility and accountability for the results. Finally, this article does not contain any studies with human participants or animals performed by any of the authors.

## References

1. Towers, G. & Chowell, G. Impact of weekday social contact patterns on the modeling of influenza transmission. J Theor Biol 312, 87–95, DOI: 10.1016/j.jtbi.2012.07.023 (2012).

2. Cooley, P. C. et al. Weekends as social distancing and their effect on the spread of influenza. Comput. Math Organ Theory 22, 71–87, DOI: 10.1007/s10588-015-9198-5 (2016).

3. Zierenberg, J. et al. New j. phys. 25 053033. New J. Phys. 25, 053033 (2023).

4. Moritz, S. et al. The risk of indoor sports and culture events for the transmission of COVID-19. Nat Commun 12, 5096, DOI: 10.1038/s41467-021-25317-9 (2021).

5. Jason, J. S. et al. Impact of weekday and weekend mobility and public policies on COVID-19 incidence and deaths across 76 large municipalities in Colombia: statistical analysis and simulation. BMC Public Heal. 22, 2460, DOI: 10.1186/s12889-022-14781-7 (2022).

6. Mossong, J. et al. Social contacts and mixing patterns relevant to the spread of infectious diseases. PLoS Medicine 5, e74, DOI: 10.1371/journal.pmed.0050074 (2008).

7. Huynh Thi Phuong et al. Changes in social contact patterns in Germany during the SARS-CoV-2 pandemic—An analysis based on the Covimod study (2025). 9th International Conference on Infectious Disease Dynamics: P2.156.

8. Chen, D. B., Wang, G. N., Zeng, A., Fu, Y. & Zhang, Y. C. Optimizing online social networks for information propagation. PLoS One 9, e96614, DOI: 10.1371/journal.pone.0096614 (2014).

9. Fernando Antonio, Atayde, J., Yamzon, M. & Sy, C. An optimization model for the design of supply chains considering disruptions from pandemic uncertainty and infection trends. Clean. Eng. Technol. 11, 100577, DOI: 10.1016/j.clet.2022.100577 (2022).

10. Javarone, M. A. Social influences in opinion dynamics: The role of conformity. Phys. A 414, 19–30, DOI: 10.1016/j.physa.2014.07.018 (2014).

11. Dike, D. O. et al. Minimization of call congestion in telecommunication system using OFDM optimization model. In 2013 IEEE International Conference on Emerging & Sustainable Technologies for Power & ICT in a Developing Society (NIGERCON), 123–128, DOI: 10.1109/NIGERCON.2013.6715646 (2013).

12. Barthe-Delanoë, A.-M., Montarnal, A., Truptil, S., Bénaben, F. & Pingaud, H. Towards the agility of collaborative workflows through an event driven approach—Application to crisis management. Int J Disaster Risk Reduct 28, 214–224, DOI: 10.1016/j.ijdrr.2018.02.029 (2018).

13. Mauras, S. et al. Mitigating COVID-19 outbreaks in workplaces and schools by hybrid telecommuting. PLOS Comput. Biol 17, e1009264, DOI: 10.1371/journal.pcbi.1009264 (2021).

14. Ponge, J. et al. Evaluating parallelization strategies for large-scale individual-based infectious disease simulations. In 2023 Winter Simulation Conference (WSC), 1088–1099, DOI: 10.1109/WSC60868.2023.10407633 (2023). See also: https://immidd.github.io/GEMS/.

15. Adamik, B. et al. Bounds on the total number of SARS-CoV-2 infections: The link between severeness rate, household attack rate and the number of undetected cases (2020).

16. Niedzielewski, K. et al. The overview, design concepts and details protocol of ICM epidemiological model (PDYN 1.5) (2022). Available at: https://ssrn.com/abstract=4039054.

17. Bicher, M. et al. Model based estimation of the SARS-CoV-2 immunization level in Austria and consequences for herd immunity effects. Sci. Reports 12, 2872, DOI: 10.1038/s41598-022-06771-x (2022).

18. Households and families. Available at: https://www.destatis.de/EN/Themes/Society-Environment/Population/Households-Families/_node.html.

19. Carrat, F. et al. Time lines of infection and disease in human influenza: A review of volunteer challenge studies. Am J Epidemiol 167, 775–785, DOI: 10.1093/aje/kwm375 (2008).

20. Ajelli, M., Poletti, P., Melegaro, A. & Merler, S. The role of different social contexts in shaping influenza transmission during the 2009 pandemic. Sci. Reports 4, 7218, DOI: 10.1038/srep07218 (2014).

21. Fadel, M. et al. Association between covid-19 infection and work exposure assessed by the mat-o-covid job exposure matrix in the constances cohort. Occup. Environ. Medicine 79, 782–789, DOI: 10.1136/oemed-2022-108436 (2022).

22. Manica, M. et al. Estimating sars-cov-2 transmission in educational settings: a retrospective cohort study. Influ. Other Respir. Viruses 17, e13049, DOI: 10.1111/irv.13049 (2023).

23. Diekmann, O., Heesterbeek, J. A. P. & Metz, J. A. J. On the definition and the computation of the basic reproduction ratio r_0 in models for infectious diseases in heterogeneous populations. J. Math. Biol. 28, 365–382 (1990).

24. Bollobás, B. Random Graphs, vol. 73 of Cambridge Studies in Advanced Mathematics (Cambridge University Press, 2001), 2 edn.

25. Hethcote, H. W. Qualitative analyses of communicable disease models. Math. Biosci. 28, 335–356, DOI: 10.1016/0025-5564(76)90132-2 (1976).

26. Prem, K., Cook, A. R. & Jit, M. Projecting social contact matrices in 152 countries using contact surveys and demographic data. PLoS Comput. Biol. 13, e1005697, DOI: 10.1371/journal.pcbi.1005697 (2017).

27. Beale, S. et al. Workplace contact patterns in England during the COVID-19 pandemic: Analysis of the Virus Watch prospective cohort study. Lancet Reg. Heal. - Eur. 16, 100352, DOI: 10.1016/j.lanepe.2022.100352 (2022).

28. Hâncean, M.-G. et al. Occupations and their impact on the spreading of COVID-19 in urban communities. Sci. Reports 12, 14115, DOI: 10.1038/s41598-022-18392-5 (2022).

29. Li, P., Dai, M., Wang, Y. & Liu, Y. Estimation of under-reporting influenza cases in Hong Kong based on Bayesian hierarchical framework. Infect. Dis. Model. 10, 946–959, DOI: 10.1016/j.idm.2025.05.002 (2025).

30. Madmon, O. & Goldberg, Y. Infectious diseases: Household modeling with missing data. Epidemics 50, 100811, DOI: 10.1016/j.epidem.2024.100811 (2025).

