## Appendices 1,2,3 for "Impact of Temporal Patterns in Working Contacts on Epidemic Spread"

### Appendix 1. Table of spectral radii difference for combinations of infectious period and latent period for the 2 weekend days case

Recalculating the spectral radii difference for the possible combinations of infectious period $I$ and latent period $L$, we obtain Tab. A1. One population group is considered: working adults. We assume that each individual has $n_{hh}$= 1 contacts in household daily. The contacts on workplace $n_{wp}$ and global setting $n_{gs}$ are shown in Tab. 2 (baseline) and Tab.3 (alternative). The used infectivity values are: $p_{hh} = 0.06$, $p_{wp} = 0.03$ and $p_{gs} = 0.0132$.

| L \ I | 1 | 2 | 3 | 4 | 5 | 6 | 7 | 8 | 9 | 10 |
|---|---|---|---|---|---|---|---|---|---|---|
| 0 (7) | -0.036 | 0.003 | 0.008 | 0.005 | 0.001 | 0.005 | 0 | -0.016 | -0.021 | -0.011 |
| 1 (8) | 0.009 | 0.015 | 0.015 | 0.014 | 0.017 | 0.013 | 0 | -0.004 | 0.005 | 0.012 |
| 2 (9) | 0.009 | 0.014 | 0.011 | 0.017 | 0.017 | 0.005 | 0 | 0.007 | 0.014 | 0.016 |
| 3 (10) | 0.009 | -0.010 | 0.011 | 0.014 | 0.001 | -0.005 | 0 | 0.006 | 0.008 | 0.016 |
| 4 (11) | 0.009 | 0.014 | 0.015 | 0.005 | -0.005 | -0.007 | 0 | 0.006 | 0.014 | 0.012 |
| 5 (12) | 0.009 | 0.015 | 0.008 | -0.019 | -0.022 | -0.007 | 0 | 0.007 | 0.005 | -0.011 |
| 6 (13) | 0.009 | 0.003 | -0.054 | -0.019 | -0.005 | -0.005 | 0 | -0.004 | -0.021 | -0.029 |

**Table A1.** Spectral radii difference $\rho_{baseline} - \rho_{alternative}$ for combinations of infectious period $I$ and latent period $L$ in case of 2 weekend days

### Appendix 2. Table of spectral radii difference for combinations of infectious period and latent period for the 3 weekend days case

Recalculating the spectral radii difference for the possible combinations of infectious period $I$ and latent period $L$, we obtain Tab. A2. One population group is considered: working adults. We assume that each individual has $n_{hh}$= 1 contacts in household daily. The contacts on workplace $n_{wp}$ and global setting $n_{gs}$ are shown in Tab. 2 (baseline) and Tab. 7 (alternative). The used infectivity values are: $p_{hh} = 0.06$, $p_{wp} = 0.03$ and $p_{gs} = 0.0132$.

| L \ I | 1 | 2 | 3 | 4 | 5 | 6 | 7 | 8 | 9 | 10 |
|---|---|---|---|---|---|---|---|---|---|---|
| 0 (7) | -0.068 | 0.002 | 0.010 | 0.019 | 0.016 | 0.012 | 0 | -0.031 | -0.044 | -0.029 |
| 1 (8) | 0.016 | 0.025 | 0.033 | 0.035 | 0.033 | 0.025 | 0 | -0.011 | 0.001 | 0.024 |
| 2 (9) | 0.016 | 0.025 | 0.032 | 0.034 | 0.033 | 0.012 | 0 | 0.007 | 0.028 | 0.041 |
| 3 (10) | 0.016 | 0.016 | 0.032 | 0.035 | 0.016 | 0 | 0 | 0.020 | 0.035 | 0.041 |
| 4 (11) | 0.016 | 0.025 | 0.033 | 0.019 | -0.012 | -0.022 | 0 | 0.020 | 0.028 | 0.024 |
| 5 (12) | 0.016 | 0.025 | 0.010 | -0.052 | -0.060 | -0.022 | 0 | 0.007 | 0.001 | -0.029 |
| 6 (13) | 0.016 | 0.002 | -0.112 | -0.052 | -0.012 | 0 | 0 | -0.011 | -0.044 | -0.063 |

**Table A2.** Spectral radii difference $\rho_{baseline} - \rho_{alternative}$ for combinations of infectious period $I$ and latent period $L$ in case of 3 weekend days scenario as alternative

### Appendix 3. Table of spectral radii difference for combinations of infectious period and latent period for alternating of 2 working days and 2 days off

Recalculating the spectral radii difference for the possible combinations of infectious period $I$ and latent period $L$, we obtain Tab. A3. One population group is considered: working adults. We assume that each individual has $n_{hh}$= 1 contacts in household daily. The contacts on workplace $n_{wp}$ and global setting $n_{gs}$ are shown in Tab. 2 (baseline) and Tab. 9 (alternative). The used infectivity values are: $p_{hh} = 0.06$, $p_{wp} = 0.03$ and $p_{gs} = 0.0132$.

| L \ I | 1 | 2 | 3 | 4 | 5 | 6 | 7 | 8 | 9 | 10 |
|---|---|---|---|---|---|---|---|---|---|---|
| 0 (4) | -0.090 | 0 | 0.013 | 0 | -0.046 | -0.034 | 0.005 | 0 | -0.041 | -0.036 |
| 1 (5) | 0.019 | 0.032 | 0.029 | 0 | 0.007 | 0.037 | 0.033 | 0 | 0.004 | 0.038 |
| 2 (6) | 0.019 | 0.032 | 0.013 | 0 | 0.032 | 0.037 | 0.005 | 0 | 0.034 | 0.038 |
| 3 (7) | 0.019 | 0 | -0.050 | 0 | 0.007 | -0.034 | -0.042 | 0 | 0.004 | -0.036 |

**Table A3.** Spectral radii difference $\rho_{baseline} - \rho_{alternative}$ for combinations of infectious period $I$ and latent period $L$ in case of 2 working days /2 days off scenario as alternative
